# Development of a spontaneous preterm birth predictive model using a panel of serum protein biomarkers for early pregnant women: A nested case–control study

**DOI:** 10.1101/2024.01.29.24301917

**Authors:** Shuang Liang, Yuling Chen, Tingting Jia, Ying Chang, Wen Li, Yongjun Piao, Xu Chen

**Affiliations:** Tianjin Central Hospital of Gynecology Obstetrics, Tianjin, China; Tianjin Key Laboratory of Human Development and Reproductive Regulation, Tianjin, China; School of Life Sciences, Tsinghua University; School of Medicine, Nankai University, Tianjin, China

**Keywords:** Preterm birth, Pregnancy, Prediction model, Biomarker

## Abstract

**Objective:** To develop a model based on first trimester maternal serum LC-MS/MS to predict spontaneous preterm birth (sPTB) < 37weeks.

**Methods:** A cohort of 2,053 women were enrolled in a tertiary maternity hospital in China from July 1, 2018 to January 31, 2019. In total, 110 singleton pregnancies (26 cases of sPTB and 84 controls) at 11–13^6/7^ gestational weeks were used for model development and internal validation. A total of 72 pregnancies (25 cases of sPTB and 47 controls) at 20-32 gestational weeks from an additional cohort of 2,167 women were used to evaluate the scalability of the prediction model. Maternal serum samples were collected at enrollment and analyzed by LC-MS/MS, and candidate proteins were used to develop an optimal predictive model by machine learning algorithms.

**Results:** A novel predictive panel with four proteins, including sFlt-1, MMP-8, ceruloplasmin, and SHBG, which was the most discriminative subset, was developed. The optimal model of logistic regression had an AUC of 0.934, with additional prediction of sPTB in second and third trimester (0.868 AUC). Importantly, higher-risk subjects defined by the prediction generally gave birth earlier than lower-risk subjects.

**Conclusion:** First trimester modeling based on maternal serum LC-MS/MS identifies pregnant women at risk of sPTB, which may provide utility in identifying women at risk at an early stage of pregnancy before clinical presentation to allow for earlier intervention.

## Introduction

Preterm birth (PTB) is defined as delivery before 37 weeks of gestation, and it is a major pregnancy issue that affects children[1]. According to the WHO, PTB is the leading cause of perinatal morbidity and mortality globally[2]. Moreover, the survivors among these premature neonates are at an increased risk of long-term morbidities, such as cerebral palsy, neurodevelopmental impairment, schizophrenia, and anxiety disorder[3–5], that result in a large economic burden[6, 7]. The application of interventions to prevent PTB remains challenging due to the inability to assess the true risk of preterm labor. Great effort has been made to predict PTB; however, known screening methods or risk factors, such as cervicovaginal fetal fibronectin (fFN) testing, prior history of spontaneous preterm birth (sPTB), and short cervical length, have not improved the preterm birth outcomes in clinical practice[8–10].

The predictors of sPTB are need to be further determined. According to the latest studies in molecular biology, sPTB is a heterogeneous disease with multiple etiologies, including intrauterine infection/inflammation, placental protein/hormonal disorders, matrix remodeling, and abnormal allogenic recognition [11–13]. This heterogeneity suggests that it cannot be accurately predicted by a single indicator. The widespread use of liquid chromatography-tandem mass spectrometry (LC-MS/MS) enables the detection of multiple protein biomarkers simultaneously, providing new hope for clinical prediction of the disease. Moreover, models built from proteomic data predict sPTB with higher accuracy and earlier in pregnancy than other omics models. In a Proteomic Assessment of Preterm Risk (PAPR) project, Saade et al. used mass spectrometry to assess maternal circulating proteomic profiles in the second trimester and identified two proteins related to PTB: insulin-like growth factor-binding protein 4 (IGFBP4) and sex hormone-binding globulin (SHBG)[7]; this provides feasibility for detecting predictive indicators of sPTB by MS.

PTB has been recently reported to be a primary placental disorder, which is due to angiogenesis disorders typified by shallow trophoblast invasion and deficient spiral artery conversion in early pregnancy[14, 15]. Kim et al. observed that patients delivered preterm had a greater degree of failure of transformation of the spiral arteries in the myometrial and decidual segments than women who delivered at term. Most of the current research on the prediction of PTB is based on biomarkers of second and late trimester, therefore, the fundamental role of placental dysfunction in early trimesters in PTB cannot be reflected, while other ‘placenta originated disease’, such as preeclampsia, was predicted by maternal serum in early pregnancy and the model performed well. Identification of early biomarkers is necessary to develop treatment strategies that reduce the impact of prematurity.

Here, we performed a proteomic profile with LC-MS/MS at the 11-13^6/7^ gestational weeks based on 44 selected candidate biomarkers found to be involved in the pathological processes of sPTB and developed a predictive model with four proteins, including soluble fms-like tyrosine kinase-1, matrix metalloproteinase-8, ceruloplasmin, and sex hormone-binding globulin, which had a distinguishing performance. Then, an additional evaluation was proceeded in women at 20–32 gestational weeks to evaluate the scalability of the model across different gestational weeks. This study provides a new panel and predictive model for sPTB and provides the potential to improve treatment and prognosis for clinical practice.

## Materials and Methods

### Ethics

This study was approved by the Ethics Committee of Tianjin Central Hospital of Obstetrics and Gynecology (2020KY023, April 2, 2020).

### Subjects

A prior development nested case–control study was conducted in a cohort of 2,053 women enrolled between January 1, 2018, and January 31, 2019, at Tianjin Central Hospital of Obstetrics and Gynecology. Subjects were included in this study after written consent was obtained from them. Healthy pregnant women between the ages of 20 and 40 years who presented to Tianjin Central Hospital of Obstetrics and Gynecology for delivery were eligible. The development cohort was based on singleton pregnancies at the time of prenatal aneuploidy serum screening at 11–13^6/7^ gestational weeks. An additional cohort of 2,167 women were used to evaluate the scalability of the prediction model. The sample size of case and control group was calculated in a ratio of 1:3 in the development cohort and 1:2 in the scalability cohort, respectively. Gestational age was determined based on the first day of the last menstrual period and was corroborated by ultrasound dating. Maternal sociodemographic data were recorded by trained healthcare staff. BMI was derived from the patient’s height and self-reported prepregnancy weight. Following delivery, data were collected for maternal and infant outcomes and complications. All deliveries were classified as term (≥37 gestational weeks), sPTB (including preterm premature rupture of membranes [PPROM]), or medically indicated preterm births (miPTB). All sPTB cases and controls in this study were individually adjudicated by the chief medical officer, and discrepancies were clarified with the principal investigator at the clinical site. With the exception of the director of clinical operations (DCO), all laboratory and data analysis personnel were blinded to the clinical data.

### Sample collection

Maternal whole blood was placed in a 4°C refrigerator for no more than 2 hours until centrifugation (2200 ⅹ g at 4°C for 10 min). After a 10-minute room temperature clotting period, 0.5-mL serum aliquots were carefully separated and stored at −80°C until subsequent analysis.

### Sample preparation

An extensive literature review was performed, and from this literature, 44 candidate biomarkers that have been found to be associated with the pathological processes implicated in PTB were selected. In Supplemental Table 1, the candidate biomarkers and molecular pathways are categorized along with the corresponding literature reviews that highlight the role of each biomarker in the pathological processes implicated in PTB. The corresponding unique peptides used for target protein identification based on MS are listed in Supplemental Table 2. All of these peptides were synthesized and purified to above 90% purity. For protein digestion of serum samples, 140 μL of 8 M urea in PBS buffer was added to 2 μL serum, followed by TCEP reduction and chloroacetamide alkylation. Then, trypsin was added at a 1:50 ratio (micrograms of enzyme to micrograms of protein). Digestion was performed at 37 ° C overnight. Peptides were acidified to a final concentration of 0.1% trifluoro-acetic acid (TFA) and desalted by tC18 Sep-Pak columns (Waters). The elution was completely dried using a SpeedVac centrifuge at 45°C, and peptides were suspended in 100 mM TEAB buffer. The synthesized peptides were also dissolved in 100 mM TEAB buffer at an appropriate concentration, reduced by TCEP and alkylated by chloroacetamide. Synthesized peptides and serum peptides were labeled by TMT6plex reagents. Each TMT group contained TMT126-labeled synthesized peptides and four serum samples labeled with TMT128-131 reagents. After quenching the labeling reaction by 5% hydroxylamine, different TMT-labeled samples were mixed together and desalted by homemade C18 StageTips. The collection material was dried and suspended in 0.1% formic acid for MS analysis.

### LC-MS/MS Analysis

Details of LC-MS/MS profiling procedures were listed in the Supplemental Methods.

### Outcomes and measurements

The outcome measures for this study were sPTB <37 weeks’ gestation.

### Data preprocessing and marker selection

Protein markers with more than 20% missing values were excluded from the analysis. The missing values of the remaining markers were imputed with mean values using the imputeTS R package. Then, a feature selection model was used to reduce the number of markers. Feature selection is a machine learning technique that is used to remove irrelevant features for prediction tasks. Feature selection approaches can be broadly divided into two types: filters and wrappers. The filters employ independent metrics to evaluate candidate feature subsets, while the wrappers use machine learning algorithms to determine the best subset. The filters are faster than wrappers, but the interaction with the final prediction model is ignored. Wrappers can identify the most discriminative subsets for the prediction model, but they are computationally expensive. Since the dimensionality (the number of protein markers) of our data was not very high, we used the wrapper approach with sequential forward selection (SFS) to select relevant markers for sPTB prediction. Starting with the empty set, the selection model added one marker at a time and repeated the procedure until the best marker subset was obtained. A support vector machine was used as an evaluation algorithm for candidate subsets generated from each round of iteration, and the 5-fold cross validation accuracy was used as a selection criterion.

### PTB prediction model development and scalability evaluation

Logistic regression (LR), support vector machine (SVM), and random forest (RF) were used to construct predictive models. To perform the internal model validation, we used 5-fold cross validation. In the 5-fold cross validation, the data were randomly divided into 5 pieces. Four pieces were used for the training, and the remaining one was used for the testing. Model discrimination was assessed by accuracy, sensitivity, specificity, precision, F-measure, and the area under the receiver operating characteristic curve (AUC), and the calibration was assessed by the calibration plot. To evaluated scalability of the model, an independent cohort consisted of different gestational weeks (20-32) were selected for LC/MS-MS analysis. Then, we also evaluated the model performances in terms of discrimination and calibration as did in development dataset. Scikit-learn python machine learning package (v1.2.0) was used for model development and scalability evaluation.

## Results

### Clinical characteristics

A total of 2,053 subjects treated at Tianjin Central Hospital of Obstetrics and Gynecology between January 1, 2018, and January 31, 2019, were enrolled in the development phase (Figure 1a). Forty-two subjects (2.05%) were excluded due to abortion or termination of pregnancy, 16 subjects (0.78%) provided insufficient information, and 377 (18.36%) were lost to follow-up. Of the remaining 1,618 subjects, 75 of them were PTBs (4.63%), including 26 (34.67%) spontaneous and 49 (65.33%) medically indicated PTBs. 84 controls were randomly selected from 1,543 participants with normal delivery. There were no statistically significant differences between the case and control groups in terms of maternal age, educational level, or sampling time (Table 1), with the exception that subjects with sPTB were more likely to have had 1 or more previous PTBs and more likely to have conceived the current pregnancy through assisted reproduction (*P*=0.012). A total of 25 cases of sPTB and 47 controls from an additional cohort of 2,167 subjects were enrolled in the scalability evaluation phase (Figure 1b). For 3 of the scalability cohort samples were degraded, so there were only 47 controls in the scalability cohort. The characteristics of the PTB cases and controls selected for the overall scalability evaluation cohort were not significantly different. The characteristics of the subjects in it are summarized in Supplemental Table 3.

**Figure 1.**
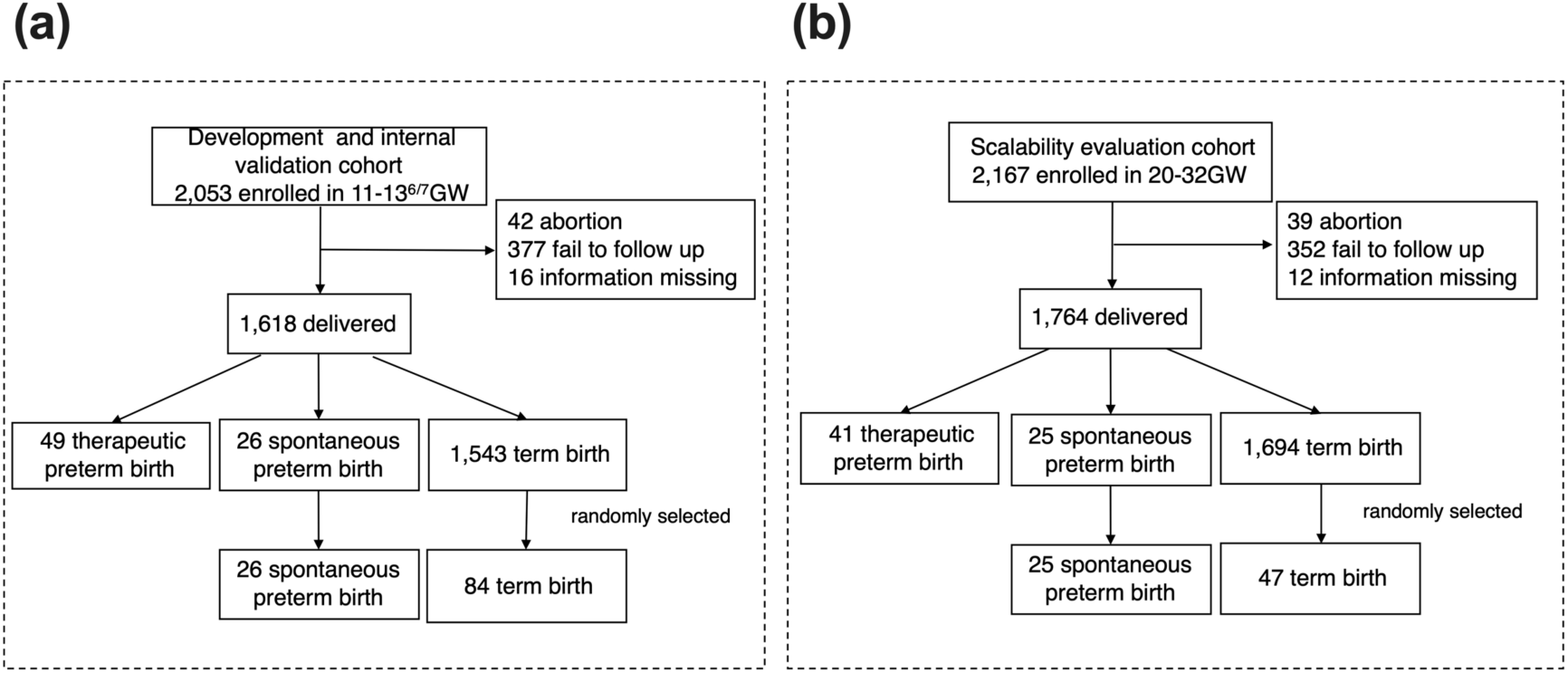
Workflow of subject enrollment for the (a) development and (b) scalability evaluation phases.

**Table 1.** Demographic characteristics of the participants.

|  | Preterm (n=26) | Term (n=84) |
| --- | --- | --- |
| Age | 31.308±0.531 | 30.143±0.431 |
| Prepregnancy BMI |  |  |
| <18.5 | 4 (15.4%) | 15 (17.9%) |
| 18.5–24.9 | 19 (73.1%) | 57 (67.9%) |
| ≥25 | 3 (11.5%) | 12 (14.3%) |
| Height | 162.577±0.633 | 164±0.588 |
| Educational level |  |  |
| <College | 3 (11.5%) | 10 (11.9%) |
| =College | 21 (80.8%) | 67 (79.8%) |
| >College | 2 (7.7%) | 7 (8.3%) |
| Cigarette smoking | 1 (0.0%) | 1 (1.2%) |
| Multigravida | 6 (23.1%) | 21 (25.0%) |
| Preterm delivery history | 3 (11.5%) | 1 (1.1%) |
| Term delivery history | 3 (11.5%) | 20 (23.8%) |
| Primigravida | 20 (76.9%) | 63 (75.0%) |
| Assisted reproduction | 3 (11.8%) | 0 (0.0%) |
| Gestational days at serum collection | 88.923±0.764 | 88.345±0.458 |
| Bleeding during pregnancy before 12 wk | 8 (30.8%) | 14 (16.7%) |

### Model development and internal validation

Of the 44 candidate biomarkers involved in the pathological processes of sPTB, 35 were detected in the maternal serum samples by LC-MS/MS analysis. Ten of the 35 proteins were detected had > 20% missing values; thus those 10 proteins were removed. Of the remaining proteins, 2 of them (INFA13 and IL5) had missing values in 8 samples, and these missing values were imputed with mean. After processing of missing values, 25 proteins were finally used for downstream analysis, and the heatmap of the data is shown in Figure 2a. The most discriminative markers soluble fms-like tyrosine kinase-1 (sFlt-1), matrix metalloproteinase-8 (MMP-8), ceruloplasmin, and sex hormone-binding globulin (SHBG) were selected as final sPTB prediction markers based on the feature selection approach. Then, predictive models were constructed using machine learning algorithms based on those 4 markers, and 5-fold cross validation was performed to evaluated the models (Figure 2b). LR were found to result in best performance with a classification accuracy of 0.918, followed by lower accuracy values with SVM (0.909) and RF (0.873) (Table 2). The average AUCs of 0.94, 0.93, and 0.92 were achieved in the 5-fold cross validation for LR, SVM, and RF, respectively (Figure 2c). Model calibration plot is shown in Figure 2d. The subjects were then divided into high or low risk group according to a predictive probability cutoff which is 2 x average sPTB rate in China (7.3%x2=14.6%), and Kaplan-Meier analysis was then conducted between these two groups (Figure 2e). It is observed that, high-risk group delivered earlier than low-risk group (*P*=0.002).

**Figure 2.**
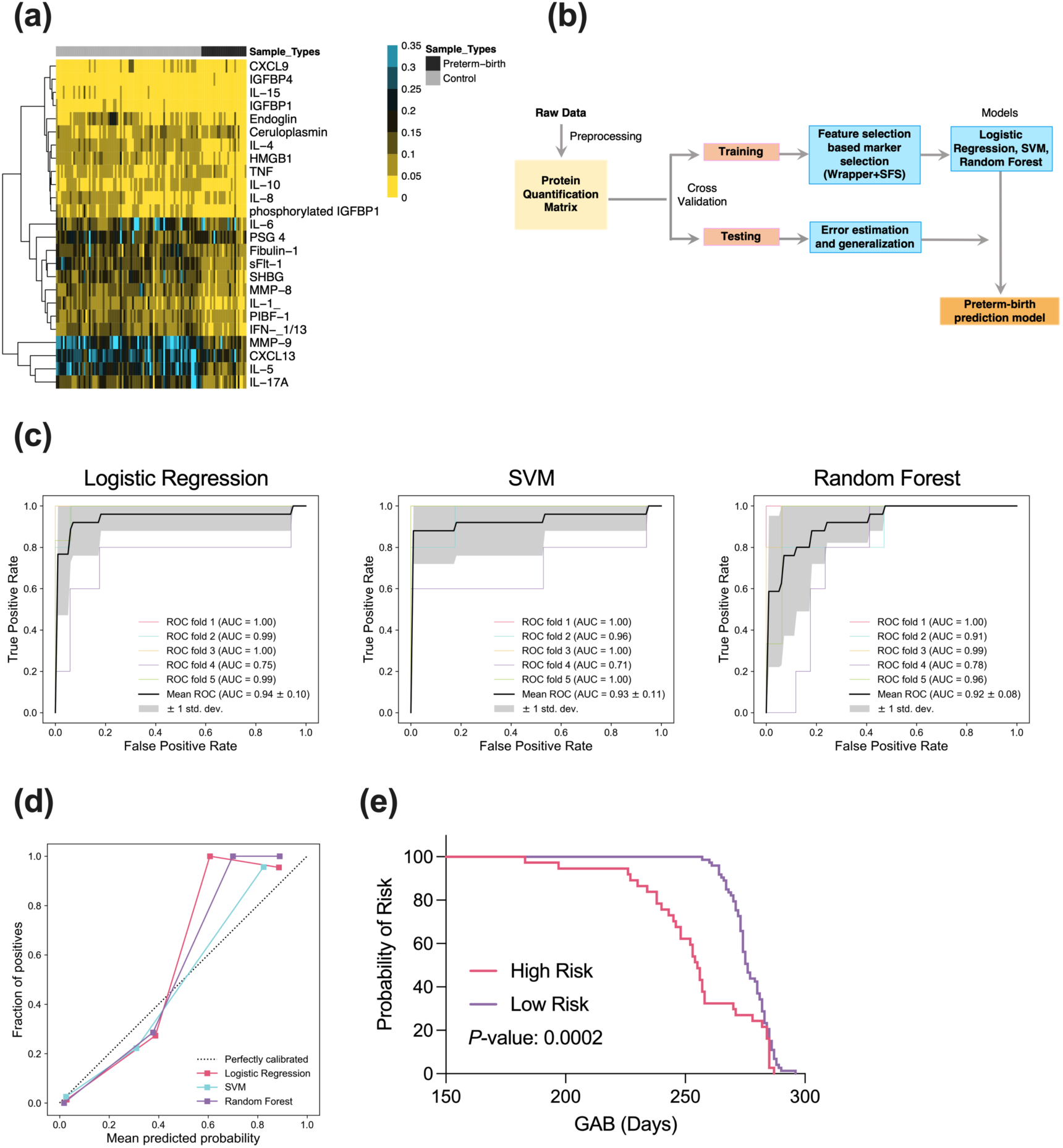
sPTB prediction model development. (a) Unsupervised clustering analysis of protein markers and between the sPTB and control groups in the training dataset. (b) Workflow for the development and validation of the prediction model. (c) ROC analysis of three different machine learning models 5-fold cross validation. (d) Calibration plot of three prediction models. (e) Kaplan-Meier analysis of high and low risk groups.

**Table 2.** Performance estimation of preterm birth prediction model in terms of classification accuracy, sensitivity, specificity, precision, and F-measure on 5-fold cross.

| Models | Acc | Sen | Spe | Precision | F-measure |
| --- | --- | --- | --- | --- | --- |
| **LR** | 0.918 | 0.767 | 0.965 | 0.887 | 0.811 |
| **SVM** | 0.909 | 0.880 | 0.918 | 0.785 | 0.823 |
| **RF** | 0.873 | 0.627 | 0.953 | 0.810 | 0.682 |

\*Acc: Accuracy, Sen: Sensitivity, Spe: Specificity, LR: Logistic regression, SVM: Support vector machine, RF: Random forest

### Scalability of the model across different gestational weeks

To further evaluate whether the prediction model is expandable for prediction of sPTB in second and third trimester, the developed model was evaluated on an independent cohort consisted of different gestational weeks (20-32). In the scalability evaluation set, LR still achieved the best performance, followed by RF and SVM (Supplemental Table 4). and ROC curves are shown in Figure 3. As shown in the figure, LR achieved the best performance with an AUC=0.868 (95% CI 0.770-0.996), followed by SVM (0.826, 0.711-0.940) and RF (0.788, 0.658-0.898), respectively. Model calibration plot is shown in Supplemental Figure 1.

**Figure 3.**
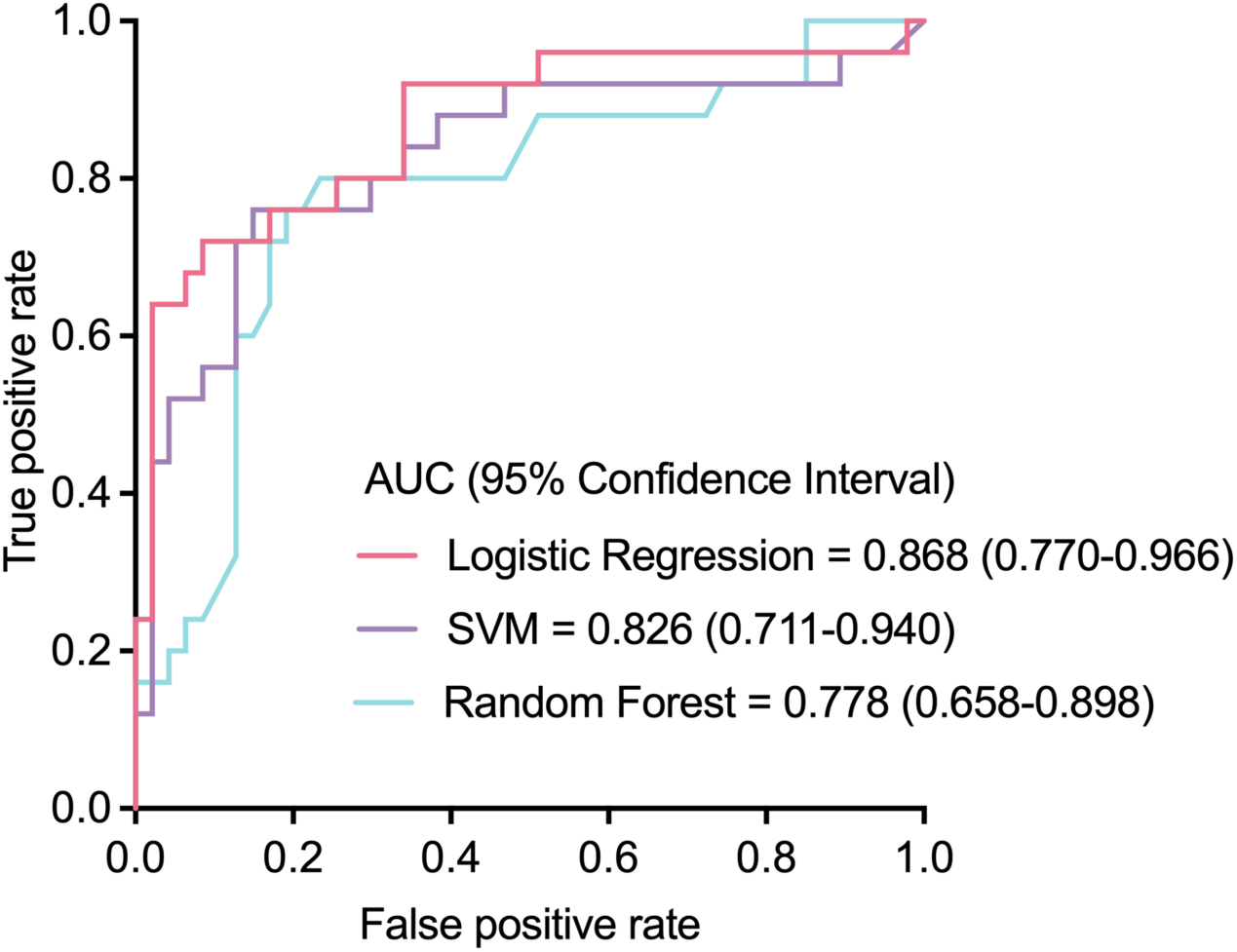
ROC analysis of three prediction models on a scalability evaluation set.

## Discussion

In this study, we developed a novel sPTB predictive panel with four proteins, including sFlt-1, MMP-8, ceruloplasmin, and SHBG, at 11-13^6/7^ gestational weeks from 44 molecules that were found to be associated with pathological processes of PTB. This model trained with logistic regression could accurately distinguish sPTB from normal deliveries with AUC of 0.94 on 5-fold cross validation, and it still worked well in an independent set of 20-32 gestational weeks with an AUC of 0.868. These 4 biomarkers were first combined to predict sPTB, and the model showed good and stable performance. This early detection may have the potential to improve treatment and prognosis for clinical practice.

Previous studies have documented several indicators for prediction of sPTB, but the prediction of sPTB still remains a challenge. A prior history of sPTD is considered to be the best measure of clinical risk to date[7]; however, more than 50% of PTBs occur in low-risk groups[21]. Cervicovaginal fetal fibronectin (fFN) can be a useful biomarker for predicting PTB within 7 to 14 days in women with contractions and mild acute preterm labor, but the predictive value of fFN for PTB more than 14 days after testing is poor[18]. It has been reported that using combinatorial biomarkers for PTB prediction had more predictive value than a single biomarker due to the heterogeneity of PTB pathology[22].

PTB was reported to be rooted in aberrant placentation at the end of the first trimester. Classically, trophoblast cells invade into the decidual and myometrial segments of the spiral arteries, and this physiologic transformation of the spiral arteries leads to substantial dilation of these vessels, which is considered key to accommodating the increased blood flow to the uteroplacental circulation[23–25]. Accumulative observations have indicated that PTB is associated with the failure of physiologic transformation of spiral arteries, which is considered canonical in the setting of PE[26–28]. The rate of failure of physiologic transformation of the myometrial segment of the spiral arteries was significantly higher for patients with PTB than for those who had a term delivery (30.9% vs. 13.6%, p=0.004)[29]. Our model based on the sampling window of 11-13^6/7^ gestational weeks, which coincided with the timing of routine obstetrical visits, enables early detection of women at risk of sPTB.

To our knowledge, this is the first time that sFlt-1, which is related to abnormal placentation, is used as one of the factors to predict sPTB. sFlt-1 is an antiangiogenic substance secreted by trophoblasts, endothelial cells and monocytes and has been proposed to be responsible for an antiangiogenic state and systemic endothelial dysfunction[30]. The concept of “placental hypoperfusion” has recently been proposed and manifests as a series of pregnancy-related complications caused by vascular remodeling disorders, which can be predicted by sFlt-1 in early pregnancy, including preeclampsia and PTB[11]. Previous studies have reported that sFlt-1 levels were higher in cases of PTB. Our study supports these findings; furthermore, our multivariate predictive model suggests that sFlt-1 may have predictive value for this group of patients.

Intra amniotic infection/inflammation and placental hormonal disorders have been associated with PTB[11]. Serum ceruloplasmin is secreted by the liver and is known to be an acute phase reactant, which modulates inflammatory responses. It was reported to be related to premature rupture of the membranes and PTB. Matrix metalloproteinase-8, also known as neutrophil collagenase, is secreted by a variety of inflammatory cells. MMP-8 is present at the initial stages of the inflammatory process and has been reported to play a role in PTB. SHBG is secreted by the liver and placenta, and together with IGFBP4, has been reported to be a predictor of PTB, as SHBG controls androgen and estrogen actions in the placental-fetal unit in response to upstream inflammatory signals[7]. Therefore, our panel includes multiple mechanisms of PTB, which may be more valuable for predicting PTB due to the heterogeneity of PTB.

PTB is a multi-mechanism disease and our panel includes molecules involved in different mechanisms of PTB with stable performances, indicating that the prediction of PTB should be based on multiple perspectives. The early detection may have the potential to improve prognosis for clinical practice on the base of additional surveillance and interventions (corticosteroids and magnesium sulfate) which have well-established benefits for newborns. Large-sample validation research in the future and would enable earlier prediction.

There are several strengths in the study. First, we chose a multi-index prediction model related to the 3 major causes of PTB (vascular remodeling disorders, intra-amniotic infection and placental hormonal disorders), which was in line with the heterogeneity of PTB. Second, we used early pregnancy as the modeling time window based on the placentation abnormalities occurring in the early stage. The early prediction may provide more time for future intervention as the PE prediction did. Third, we used an independent set consisted of 20-32 gestational weeks to evaluate its scalability, which showed that our model could be applicable for various period of gestation.

There are still limitations in the present study. First, our study was a retrospective nested case-control study; therefore, mixed factors cannot be well controlled. Second, the participants were exclusively from a single center and the dataset was small, further optimization of the model requires large-sample, multicenter validation. Third, this study was not stratified to assess these markers for sPTB at <32weeks’ gestation, which was very meaningful for the prognosis for newborns, due to the limited number of sPTB cases.

## Trial registration

Chinese Clinical Trial Registry, ChiCTR2100052202, Date of Registration: 2018-01-01, Date of initial participant enrollment: 2018-07-01, http://www.chictr.org.cn/enindex.aspx.

## Data Availability

All data produced in the present study are available upon reasonable request to the authors.

