## Supplemental Table 1 for "Development of a spontaneous preterm birth predictive model using a panel of serum protein biomarkers for early pregnant women: A nested case–control study"

### Supplemental Methods

#### LC-MS/MS Analysis

For model establishment, peptides were loaded onto a trap column (75 μm×20 mm, 3 μm C18,100 Å, 164535, Thermo Fisher Scientific) with a maximum pressure of 600 bar using mobile phase A (0.1% formic acid in H2O), separated on an analytical column (75 μm×150 mm, 3 μm C18,100 Å, 164568, Thermo Fisher Scientific) with a gradient of 5-55% mobile phase B (80% acetonitrile and 0.08% formic acid) at a flow rate of 300 nL/min for 130 min. A Q-Exactive HF-X mass spectrometer was programmed to acquire PRM combined with a full scan. Full scan acquisition was performed with a resolution of 60,000 at 200 m/z ($3\times10^6$ ions accumulated with a maximum injection time of 200 ms) to cover the scan range of 350-2000 m/z. PRM acquisition was performed using a resolution of 30,000 at 200 m/z, isolation windows of 0.4 m/z, target AGC values of $2\times10^5$, and a maximum injection time of 120 ms. Fragmentation was performed with a normalized collision energy of 32 by HCD (high-energy collision dissociation).

For model validation, peptides were loaded and separated by the same HPLC system with a 120-min gradient. An Orbitrap Fusion Lumos Tribrid mass spectrometer was programmed in ms2 and ms3 PRM combined with a full scan. Full scan acquisition was performed with a resolution of 120,000 at 200 m/z ($4\times10^5$ ions accumulated with a maximum injection time of 50 ms) to cover the scan range of 350-1550 m/z. ms2 analysis was performed for the target m/z by using a resolution of 15,000 at 200 m/z, isolation windows of 0.7 m/z, target AGC values of $5\times10^4$, and a maximum injection time of 22 ms. Collision-induced dissociation (CID) fragmentation was performed with a normalized collision energy of 35. Target ms3 analysis was performed using a resolution of 50,000 at 200 m/z, ms2 isolation windows of 3 m/z, target AGC values of $5\times10^4$, and a maximum injection time of 86 ms. HCD fragmentation was performed with a normalized collision energy of 38.

The MS/MS data were searched against target peptide sequences using the SEQUEST search engine in Proteome Discoverer 2.3 software (PD2.3). The search criteria were as follows: full tryptic specificity was required; no missed cleavages were allowed; carbamidomethylation (C) and TMT6plex (K and N-terminal) were set as the fixed modifications; phosphorylation (S/T/Y) was set as the variable modification; precursor ion mass tolerances were set at 10 ppm for all MS acquired in an Orbitrap mass analyzer; and fragment ion mass tolerance was set at 20 mmu for all MS2 spectra acquired. Relative peptide quantification compared with the synthesized peptide standard was performed using PD2.3 based on the intensities of five reporter ions per peptide in the ms2 or ms3 spectra.

### Supplemental Table1. Biological process associated with candidate biomarkers

| Biological pathway | Candidate biomarker | Author, year, reference |
|---|---|---|
| Intrauterine infection/Inflammation | interleukin 1 alpha | Ulla-Britt Wennerholm et al,1998[1]; K. Motomura et al,2020[2] |
| | Interleukin-10 | Goldenberg RL et al,2001[3] |
| | Interleukin-15 | S. J. Fortunato et al,1998[4]; L. A. Gomez et al,2020[5] |
| | Interleukin-17A | L. A. Gomez et al,2020[5]; S. Park et al,2020[6];Yi Xu et al,2017[7] |
| | Interleukin-17F | L. A. Gomez et al,2020[5] |
| | Interleukin-6 | Eliane Moura et al,2009[8]; Inglis SR et al,1994[9]; Thomakos N et al,2010[10] |
| | Interleukin-1 beta | M. Schmid et al,2012[11]; Immaculate M et al,2016[12] |
| | Interleukin-12 subunit beta | S. El-Shazly et al,2004[13] |
| | Interleukin-2 | Curry AE et al,2009[14] |
| | Interleukin-8 | Dowd J et al,2008[15]; Sakai M et al,2004[16]; Sakai M et al,2004[17] |
| | Interleukin-5 | Tatiana Hountohotegbe et al,2020[18] |
| | Interleukin-12 subunit alpha | S. El-Shazly et al,2004[13] |
| | Interleukin-4 | V. S. Belousova et al,2019[19] |
| | High mobility group protein B1 | E. Radnaa et al,2021[20] |
| | Tumor necrosis factor | Eliane Moura et al,2009[8]; Inglis SR et al,1994[9]; Thomakos N et al,2010[10] |
| | Interferon gamma | Eliane Moura et al,2009[8]; Curry AE et al,2009[14] |
| | Alpha-2-HS-glycoprotein | Attila Molvarec et al, 2009[21] |
| | Alpha-fetoprotein | Jilin Hu, et al,2019[22], |
| | Ceruloplasmin | A. Seval Ozgu-Erdinc et al,2014[23] |
| | Colony-stimulating factor-2 | Rachel G. Sinkey et al,2020[24] |
| | C-X-C motif chemokine ligand 10 | Stefania Ronzoni et al,2018[25] |
| | transthyretin | F. J. Rosales et al, 1996[26] ; A. Myron Johnson et al,2007[27] |
| Placental protein/ hormona –related biomarker | Corticoliberin | Wei Perng et al,2020[28] |
| | Sex hormone-binding globulin | George R. Saade et al,2016[29] |
| | Progesterone-induced-blocking factor 1 | B. Huang et al,2017[30]; |
| | Insulin-like growth factor-binding protein 4 | George R. Saade et al,2016[29] |
| | Insulin-like growth factor-binding protein1 | R. Devliegeret al,2009[31]; D. Balic et al,2008[32] |
| | phI-Insulin-like growth factor-binding protein 1 | D. Paternoster et al,2009[33]; V. Wiwanitkit,2010[34]; O. Altinkaya et al,2009[35] |
| | Pregnancy-specific beta-1-glycoprotein 3 | J. Warren2018[36] |
| | Pregnancy-specific beta-1-glycoprotein 4 | J. Warren2018[36] |
| | Pregnancy associated plasma protein –A | Marja Kaijomaa et al,[37]; Alice E. Hughes et al, 2019[38] |
| | chorionic gonadotropin subunit beta 3 | Ida Kirkegaard et al,2010[39]; Gayathri Rengaraj et al,2007[40] |
| Immunity | C-X-C motif chemokine 9 | S. Ronzoni et al,2019[25] |
| | C-X-C chemokine receptor type 5 | C. Silwedel et al,2019[41] |
| | C-X-C motif chemokine 13 | Y. Luo et al,2020[42] |
| Matrix remodeling | matrix metalloproteinase 8 | Yoon BH et al,2001[43] |
| | matrix metalloproteinase 9 | H. J. Kim et al,2020[44]; J. W. Park et al,2019[45] |

| | | |
|---|---|---|
| | Fibulin-1 | Satoko Ito et al,2020[46] |
| | vitronectin | Kimie Date et al, 2019[47]; Teresa Cobo et al, 2018[48] |
| Angiogenesis disorders | Vascular endothelial growth factor receptor 1 | Sean Lim et al,2021[49]; C. Villalain et al,2020[50] |
| | Endoglin | T. Chaiworapongsa et al,2009[51]; |
| | Dickkopf-related protein 1 | SHENG-JUN JIANG et al,2015[52]; |
| | Interferon alpha-1-13 | Danieli Andrade et al,2015[53] |
| | Alpha-fetoprotein | OLIN D. LIANG et al,2004[54] |
| | placental growth factor | Mahsa Matin et al,2020[55]; A. Leanos-Miranda et al,2020[56]; John R. Barton et al,[57] |

**Supplemental Table 2. Peptide list of 44 PTB potential biomarkers**

| NO | Uniprot Accession | Gene Name | Peptide |
|---|---|---|---|
| 1 | P02771 | AFP | GYQELLEK |
| 2 | Q13219 | PAPPA | ALYFSGR |
| 3 | P17948 | FLT1 | GFIISNATYK |
| 4 | P05231 | IL6 | YILDGISALR |
| 5 | P01584 | IL1B | ISDHHYSK |
| 6 | P51911 | CNN1 | LQPGSVK |
| 7 | P06850 | CRH | MGEEYFLR |
| 8 | P60568 | IL2 | WITFCQSIISTLT |
| 9 | P10145 | CXCL8 | VIESGPHCANTEIIVK |
| 10 | P22894 | MMP8 | TVQDYLEK |
| 11 | P14780 | MMP9 | QLAEEYLYR |
| 12 | Q8WXW3 | PIBF1 | ELQLSTESK |
| 13 | P0DN86 | CGB3 | APPPSLPSPSR |
| 14 | P02765 | AHSG | EATEAAK |
| 15 | P00450 | CP | DNEDFQESNR |
| 16 | P23142 | FBLN1 | TGYYFDGISR |
| 17 | P49763 | PGF | CECRPLR |
| 18 | Q16557 | PSG3 | LFIPQITTK |
| 19 | Q00888 | PSG4 | TLFIFGVTK |
| 20 | P17813 | ENG | GEVTYTTSQVSK |
| 21 | P02766 | TTR | VLDAVR |
| 22 | P04004 | VTN | NGSLFAFR |
| 23 | P02778 | CXCL10 | CLNPESK |
| 24 | P29460 | IL12B | TLTIQVK |
| 25 | P01579 | IFNG | DDQSIQK |
| 26 | O43927 | CXCL13 | SIVCVDPQAEWIQR |
| 27 | Q07325 | CXCL9 | IEIIATLK |
| 28 | P32302 | CXCR5 | TVIALHK |
| 29 | O94907 | DKK1 | GQEGSVCLR |
| 30 | P01562 | IFNA1 | ITLYLTEK |
| 31 | P29459 | IL12A | AVSNMLQK |
| 32 | P05112 | IL4 | TLNSLTEQK |
| 33 | P05113 | IL5 | ETLALLSTHR |
| 34 | P09429 | HMGB1 | GEHPGLSIGDVAK |
| 35 | P01583 | IL1A | FDMGAYK |
| 36 | P01375 | TNF | VNLLSAIK |
| 37 | P04141 | CSF2 | LLNLSR |
| 38 | P22301 | IL10 | ESLLEDFK |
| 39 | P40933 | IL15 | ECEELEEK |

| | | | |
|---|---|---|---|
| 40 | Q16552 | IL17A | YPSVIWEAK |
| 41 | Q96PD4 | IL17F | LDIGIINENQR |
| 42 | P08833 | IGFBP1 | AQETSGEEISK |
| 43 | P08833 | IGFBP1 | AQETS(p)GEEISK |
| 44 | P04278 | SHBG | QAEISASAPTSLR |
| 45 | P22692 | IGFBP4 | QCHPALDGQR |
| 46 | P04278 | SHBG | VVLSQGSK |
| 47 | P04278 | SHBG | LDVDQALNR |
| 48 | P22692 | IGFBP4 | LPGGLEPK |

AQETS(p)GEEISK stand for that the $5^{th}$ S is phosphorylated.

**Supplemental Table 3.** Clinical characteristics of the population of scalability set

| | Preterm (n=27) | Term (n=45) |
|---|---|---|
| Age | 32.846 ±0.780 | 32.289±0.672 |
| Prepregnancy BMI | | |
| <18.5 | 2 (7.4%) | 5 (11.1%) |
| 18.5–24.9 | 19 (70.4%) | 30 (66. 7%) |
| ≥25 | 6 (22.2%) | 10(22.2%) |
| Height | 162.250±1.316 | 163.806±0.650 |
| Educational level | 2 (22.2%) | 2 (22.2%) |
| <College | 4 (14.8%) | 5(11.1%) |
| =College | 20 (74.17%) | 35 (77.8%) |
| >College | 3（11.1%） | 5（11.1%） |
| Cigarette smoking | 1 (0.0%) | 1 (0.0%) |
| Multigravida | 11 | 14 |
| Preterm delivery history | 2 (7.4%) | 2 (4.4%) |
| Term delivery history | 9（33.3%） | 12（26.6%） |
| Primigravida | 16 (59.3%) | 31 (68.9%) |
| Assisted reproduction | 4 (14.8%) | 2 (4.4%) |
| Gestational days at serum collection | 191.33±6.25 | 198.87±4.26 |
| Bleeding during pregnancy | | |
| before 12 wk | 6 (22.2% | 9 (20.2%) |

**Supplemental Table 4**. Model performances in terms of classification accuracy, sensitivity, specificity, precision, and F-measure on scalability validation set (20-32 GW).

| **Models** | Acc | Sen | Spe | Precision | F-measure |
|---|---|---|---|---|---|
| **LR** | 0.847 | 0.720 | 0.915 | 0.818 | 0.766 |
| **SVM** | 0.764 | 0.760 | 0.766 | 0.633 | 0.691 |
| **RF** | 0.792 | 0.720 | 0.830 | 0.692 | 0.706 |

*Acc: Accuracy, Sen: Sensitivity, Spe: Specificity, LR: Logistic regression, SVM: Support vector machine, RF: Random forest

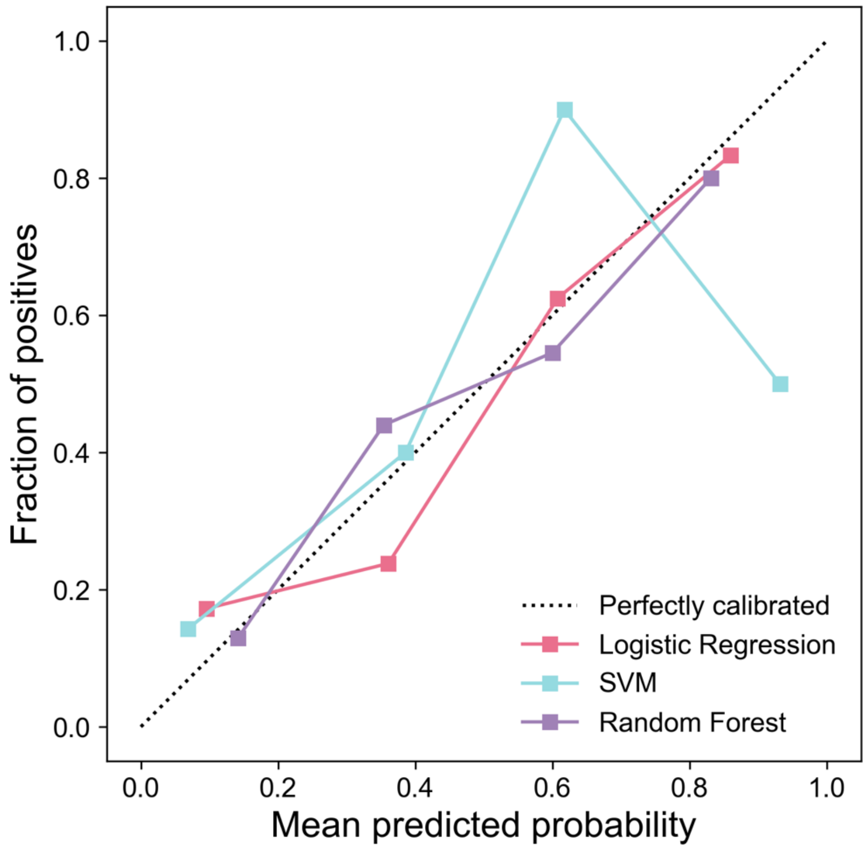


**Supplemental Figure 1**. Calibration plots of three models on scalability evaluation set (20-32 GW).
